# Respi-Radar – a tool to monitor respiratory infections in Belgium during the winter season 2023- 2024

**DOI:** 10.1101/2024.11.13.24317167

**Authors:** Géraldine De Muylder, Simon Couvreur, Giulietta Stefani, Claire Brugerolles, Yinthe Dockx, Milena Callies, Raphaël Janssens, Laurane De Mot, Nathalie Bossuyt, Jorgen Stassijns

## Abstract

Following the experience gathered during the COVID-19 crisis, the Respi-Radar was developed by the Belgian Risk Assessment Group (RAG) in the summer of 2023, with the aim of assessing the epidemiological situation of respiratory infections and informing public health preparedness and response in Belgium.

The Respi-Radar consisted of four levels (green, yellow, orange and red) which indicated the extent of viral circulation and/or pressure on the healthcare system. Depending on the level, specific measures would apply to reverse the trend. The Respi-Radar used six indicators, from the Influenza-Like Illness and Severe Acute Respiratory Infection sentinel surveillances in nursing homes, primary and secondary care, as well as from the wastewater surveillance. Additional information, such as data from the national reference laboratories, also contributed to assessing the situation.

Based on the Respi-Radar tool, the RAG regularly evaluated the epidemiological situation of respiratory infections between September 2023 and March 2024.

The Respi-Radar tool appeared useful for following trends and effectively communicating on the epidemiological situation of respiratory infections. Applying specific measures depending on a level was less straightforward. The experience gained using the Respi-Radar tool was key to determine the best approach to assess and manage the epidemiological situation for future respiratory seasons.

## 1 Introduction

In Belgium, as in other countries, the COVID-19 crisis led to the enhancement of existing surveillance systems and to the development of novel structures resulting in unprecedented amounts and types of data available. So called “management tools” were created to carry out an evaluation of the epidemiological situation of COVID-19, on a regular basis and in a structured way. The tools incorporated key indicators and thresholds to define risk levels, which would ultimately offer guidance to policy makers (1).

These tools, developed by the Risk Assessment Group (RAG, a scientific expert group coordinated by the Belgian Institute of Health, Sciensano (2)) were successively used for COVID-19 between September 2020 and August 2023, as already described (1). The last COVID-19 management tool, from December 2021, distinguished three levels: Epidemiological situation under control (Level 1), Increasing viral circulation potentially leading to pressure on the health care system (Level 2), and High viral circulation with possible health care system overload (Level 3). The levels were based on several indicators, essentially reflecting the COVID-19-related pressure on the healthcare system (number of hospital admissions due to COVID-19, COVID-19-related intensive care unit (ICU) occupancy and number of consultations of general practitioners (GP) for suspicion of COVID-19), as well as other indicators such as positivity rate for symptomatic patients, reproductive number (Rt) and 14-day incidence of cases.

In the context of co-circulation of SARS-CoV-2 and other respiratory pathogens, it became necessary to develop a management tool that would take into account not only SARS-CoV-2 but also other respiratory pathogens. The “Respi-Radar” tool was introduced in the summer 2023, with the aim of assessing the epidemiological situation of all major respiratory infections and informing public health preparedness and response.

This document first describes the Respi-Radar tool, its implementation and how it was used during the 2023-2024 respiratory season. It then presents a brief evaluation of the tool and discusses its advantages and drawbacks.

## 1 Description of the Respi-Radar tool

### a. Indicators used

The Respi-Radar was based on five pathogen-agnostic indicators from sentinel surveillance systems of respiratory infections, as well as one COVID-19-specific indicator. These indicators allowed for monitoring the circulation of respiratory pathogens, the severity of diseases and their impact on the healthcare system. They are listed in Table 1.

**Table 1.** Indicators used in the Respi-Radar.

| Indicators | Source | Reference |
| --- | --- | --- |
| Weekly incidence of consultations of general practitioners (GPs) for influenza-like illness (ILI), per 100,000 inhabitants | Sentinel network of GPs | (8) |
| Weekly incidence of consultations of GPs for other Acute Respiratory Infections (ARI), per 100,000 inhabitants | Sentinel network of GPs | (8) |
| Weekly incidence of ILI cases in nursing homes, per 1,000 nursing home residents | Sentinel network of nursing homes | (9) |
| Weekly incidence of hospitalization for Severe Acute Respiratory Infections (SARI), per 100,000 inhabitants | Sentinel network of hospitals | (8) |
| Weekly incidence of SARI hospitalization with severe complications (invasive ventilation, ECMO, admission to intensive care, , Acute Respiratory Distress Syndrom, death), per 100,000 inhabitants | Sentinel network of hospitals | (8) |
| number of wastewater treatment plants with high concentrations of SARS-CoV-2 | Wastewater surveillance | (10) |

### b. Definition of thresholds

Different thresholds were defined for each indicator. For the incidence of GP consultations for Influenza-Like Illness (ILI), the incidence of GP consultations for Acute Respiratory Infections (ARI) and the incidence of hospital admissions for Severe Acute Respiratory Infections (SARI), the levels were defined by the Moving Epidemic Method (MEM), with support from ECDC for ILI. The MEM uses historical data from past epidemic waves to identify potential upcoming epidemics and assess their intensity (4). For the incidence of ILI cases in nursing homes and the number of wastewater treatment plants with a high concentration of SARS-CoV-2, the historical data were insufficient to use the MEM. Thresholds were therefore set by experts, by consensus. All thresholds are summarized in Table 2.

**Table 2.** Risk levels in the Respi-Radar and thresholds defined for each indicator.

| Risk level | Consultations GPs for ILI* | Consultations GPs for ARI* | ILI in nursing homes** | Hospitalisations for SARI* | Severity SARI*** | SARS-CoV-2 wastewater**** |
| --- | --- | --- | --- | --- | --- | --- |
| Green | <128 | <1208 | <7 | <4,4 | <0,68 | <5 |
| Yellow | 128-507 | 1208-1293 | 7-13 | 4,4-9,8 | 0,68 - 1,4 | 5 – 10 stations + |
| Orange | 508-783 | 1294-1984 | 14-20 | 9,9-33,7 | 1,41 - 3,03 | 11-15 stations + |
| Red | >783 | >1984 | >20 | >33,7 | > 3,03 | > 15 stations + |

\* weekly incidence per 100,000 inhabitants
\*\* weekly incidence per 1,000 nursing home residents
\*\*\* weekly incidence per 100,000 inhabitants. A complication is defined as death, ARDS, admission to ICU, ECMO or invasive ventilation..
\*\*\*\* Number of treatment plants with high concentrations of SARS-CoV-2 as defined in (10)

### c. Definition of Respi-Radar levels

The Respi-Radar was divided into four levels (Table 2). The green level was the baseline and reflected a non-epidemic situation. The yellow level represented a situation where the epidemic threshold was passed for some indicators, pathogens were circulating, but the impact on the healthcare system remained limited. The orange level was characterized by a moderate pathogen circulation, which exerted some pressure on the healthcare system. The red level reflected a situation where pathogen circulation was high, with an important risk of healthcare system overload. The thresholds set for each indicator determined the Respi-Radar levels.

### d. Functioning of the Respi-Radar

Data from the surveillance of respiratory infections were available once a week and compiled in the weekly bulletin of acute respiratory infections (3).

Every week, the experts of the RAG analyzed the indicators defining the Respi-Radar and estimated an overall risk level which characterized the epidemiological situation for the week under review. The assessment was based on the indicators of the Respi-Radar but other relevant information could also be included when needed, for instance the European (through ERVISS (4)) and international situation, genomic surveillance data or qualitative information provided by the experts of the group. In addition to estimating the risk level, the RAG could propose appropriate actions, such as emphasizing the importance of vaccination and basic protective measures or reinforcing communication with relevant groups.

The risk level and actions proposed by the RAG were then presented weekly to the Risk Management Group (RMG, composed of representatives of health authorities -administration and ministries- and coordinated by the ministry of health), which validated the analysis and decided on measures to be applied (5). Specific sets of measures adapted to each risk level of the Respi-Radar were developed by various advisory bodies (6).

## 3- Use of the Respi-Radar tool during the respiratory season 2023-2024

Table 3 shows the levels of the indicators by week as well as the assessment made by the RAG, between July 2023 (week 29) and March 2024 (week 13). The epidemiological situation for respiratory infections was assessed as being at baseline (risk level *green*) between July and mid-November 2023 (weeks 29 to 45). From November 23, 2023 (week 46) to January 21, 2024 (week 3), the situation was assessed as being at risk level *yellow*, because several indicators showed increasing trends. Nevertheless, the situation was under control with a limited impact on healthcare (first and second line). The increase was due to a rise in RSV circulation in October, followed by increased SARS-CoV-2 circulation in December. From January 22, 2024 (week 4) to February 18, 2024 (week 7), the epidemiological situation for respiratory infections was at risk level *orange*, mainly due to the increased circulation of Influenza and the subsequent pressure on healthcare. The epidemiological situation was assessed as being at risk level *yellow* again on February 19, 2024 (week 8), because the impact on the healthcare system was limited, despite viral circulation above the epidemic threshold,. From March 11, 2024 (week 11) and until the end of the respiratory season, the epidemiological situation for respiratory infections remained at baseline.

**Table 3.** Respi Radar evaluation July 2023 – March 2024.

| Week | Indicators acute respiratory infections |  |  |  |  | COVID-19 specific indicator | Evaluation RAG |
| --- | --- | --- | --- | --- | --- | --- | --- |
|  | Consultations GPs for ILI symptoms* | Consultations GPs for ARI * | ILI in nursing homes ** | Hospital admissions for SARI* | SARI hospitalizations with severe complications**<br>* | Concentration SARS-CoV-2 in wastewater**** |  |
| 2023w29 | 30 | 353 | 3 | 1,3 | 0,5 | 0 | green |
| 2023w30 | 37 | 401 | 3 | 2,3 | 0,1 | 0 | green |
| 2023w31 | 59 | 419 | 3 | 2,9 | 0,7 | 3 | green |
| 2023w32 | 29 | 387 | 2 | 2 | 0,3 | 7 | green |
| 2023w33 | 79 | 383 | 3 | 3,9 | 0,6 | 5 | green |
| 2023w34 | 87 | 447 | 5 | 5,1 | 0,7 | 8 | green |
| 2023w35 | 82 | 526 | 7 | 6,1 | 0,5 | 10 | green |
| 2023w36 | 150 | 667 | 6 | 6,4 | 0 | 14 | green |
| 2023w37 | 138 | 650 | 4 | 4,9 | 0 | 7 | green |
| 2023w38 | 142 | 774 | 5 | 6,2 | 0 | 9 | green |
| 2023w39 | 181 | 994 | 8 | 8,2 | 0,4 | 14 | green |
| 2023w40 | 149 | 1105 | 7 | 6,6 | 0,4 | 10 | green |
| 2023w41 | 153 | 1074 | 4 | 6,7 | 0,3 | 15 | green |
| 2023w42 | 159 | 1129 | 6 | 8,5 | 0,4 | 11 | green |
| 2023w43 | 68 | 938 | 7 | 7,3 | 1,2 | 8 | green |
| 2023w44 | 159 | 906 | 8 | 11,3 | 0,7 | 9 | green |
| 2023w45 | 191 | 1367 | 5 | 12,1 | 0,3 | 10 | green |
| 2023w46 | 184 | 1087 | 13 | 11,1 | 2,7 | 14 | yellow |
| 2023w47 | 225 | 1169 | 10 | 12,7 | 1,8 | 19 | yellow |
| 2023w48 | 242 | 1330 | 11 | 13,6 | 1,3 | 20 | yellow |
| 2023w49 | 259 | 1487 | 15 | 12,6 | 2,2 | 24 | yellow |
| 2023w50 | 399 | 1426 | 5 | 12,6 | 2,2 | 22 | yellow |
| 2023w51 | 450 | 1518 | 9 | 16,7 | 1,3 | 27 | yellow |
| 2023w52 | 167 | 1212 | 8 | 14,5 | 1,4 | 19 | yellow |
| 2024w01 | 216 | 971 | 11 | 15,4 | 1,8 | 11 | yellow |
| 2024w02 | 298 | 1059 | 13 | 13,9 | 2 | 11 | yellow |
| 2024w03 | 325 | 1200 | 13 | 11,2 | 1,4 | 13 | yellow |
| 2024w04 | 636 | 1444 | 11 | 12,5 | 2,7 | 11 | orange |
| 2024w05 | 588 | 1623 | 12 | 12,9 | 1,0 | 9 | orange |
| 2024w06 | 557 | 1252 | 16 | 14,8 | 1,5 | 3 | orange |
| 2024w07 | 289 | 1061 | 7 | 15,9 | 1,5 | 4 | orange |
| 2024w08 | 333 | 1205 | 6 | 11,6 | 0,7 | 6 | yellow |
| 2024w09 | 228 | 921 | 8 | 11,5 | 0,9 | 4 | yellow |
| 2024w10 | 127 | 975 | 7 | 9,1 | 0,6 | 1 | yellow |
| 2024w11 | 153 | 813 | 8 | 9,2 | 1,5 | 2 | green |
| 2024w12 | 76 | 865 | 4 | 9,9 | 0,9 | 0 | green |
| 2024w13 | 95 | 940 | 3 | 8,4 | 1,1 | 0 | green |

In accordance with the estimated risk level, the RAG proposed several actions over time. In September 2023, the risk level was *green*, but in light of the upcoming respiratory season, the RAG recommended that the existing guidelines for managing cases of COVID-19 and respiratory infections in general would be repeated. In October 2023, still at risk level *green*, the RAG stressed the importance of maintaining basic protective measures (washing hands, staying at home or wearing a mask in case of symptoms). The RAG also emphasized the importance of vaccination against respiratory pathogens (Influenza, SARS-CoV-2, RSV and pneumococcus) for vulnerable populations. In November 2023, when the risk level was increased to *yellow*, the RAG referred to the recommendations formulated by other advisory bodies for the general population in Belgium: stay at home in case of symptoms, ventilate indoor spaces, wear a mask in case of symptoms, and vaccinate at-risk populations against respiratory pathogens. Recommendations for the healthcare sector were proposed by the RMG. In January 2024, when the risk level was raised to *orange*, no additional measures were set, but the RMG reinforced communication towards the public and the healthcare sector to reiterate the measures described above and stress the importance of ventilation and protection of vulnerable people.

## 4- Evaluation of the Respi-Radar

In the spring of 2024, the RAG conducted an evaluation of the Respi-Radar which included the analysis of the relevance and sensitivity of the chosen indicators, the timely availability of data, the adequacy of the thresholds and the actions taken. The evaluation was based on two components: (i) an analysis of the evolution of each indicator monitored within the Respi-Radar during the 2023-2024 season and their alignment with the global level defined each week, and (ii) a survey carried out among the experts participating to the RAG.

### a. Analysis of the indicators of the Respi-Radar

First, the completeness of the data available for the Respi-Radar evaluation was assessed for each indicator by comparing the data available at the time of the evaluation and the data when consolidated (Figure 1).

**Figure 1.**
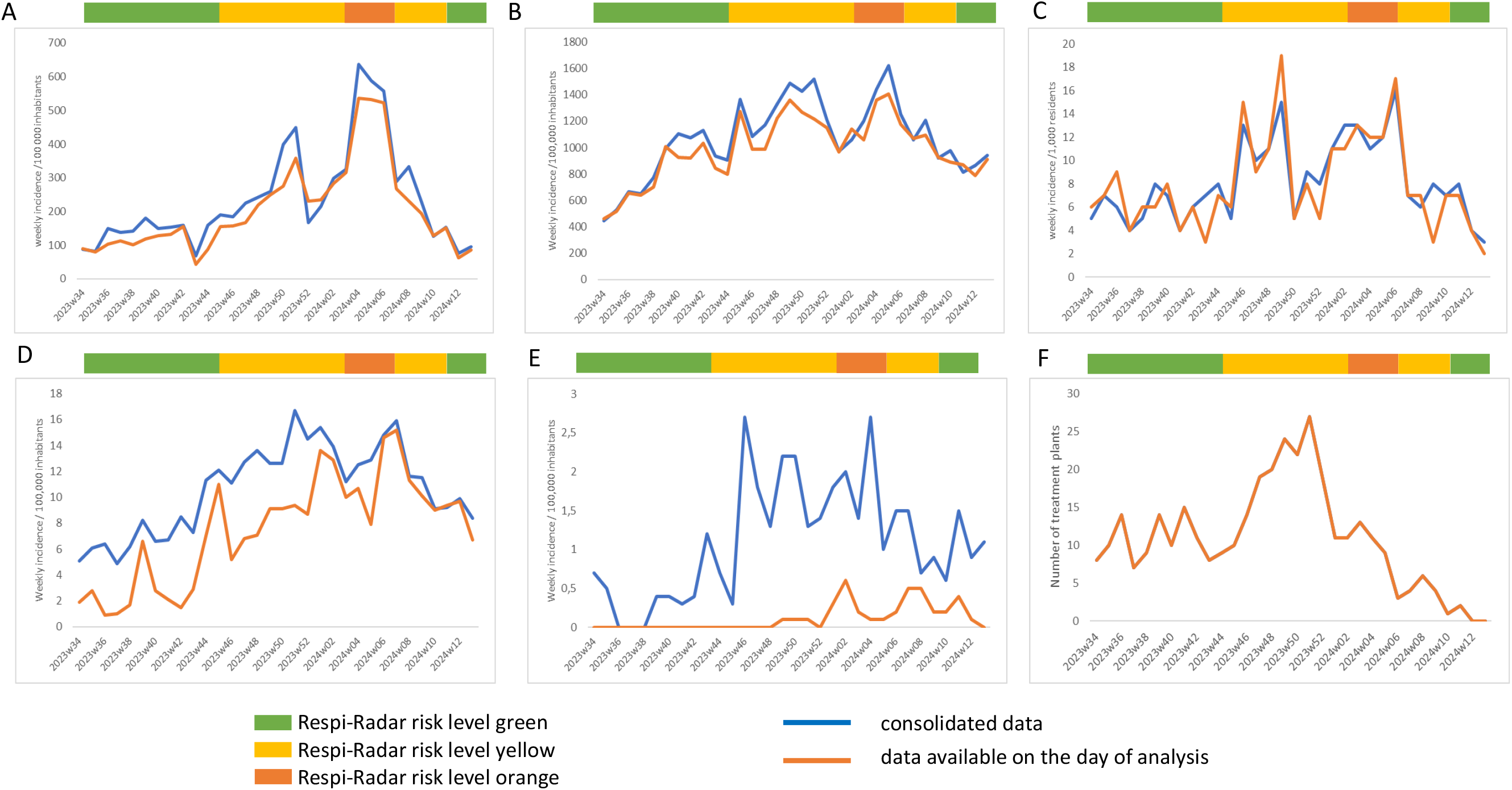
Analysis of the indicators used in the Respi-Radar. Data available at the time of the Respi Radar analysis (orange line) were compared to the same data when consolidated (blue line) from week 34 2023 to week 13 2024. Colored boxes (green, yellow or orange) indicate the overall risk level defined weekly through the Respi-Radar. A-Weekly incidence of consultations at GP practices for ILI symptoms; B-Weekly incidence of consultations at GP practices for ARI; C-Weekly incidence of ILI in nursing homes; D-Weekly incidence of hospital admissions for SARI; E-Weekly incidence of SARI hospitalizations with severe complications; F-Number of treatment plants with high concentrations of SARS-CoV-2.

Data from the sentinel networks of GPs and nursing homes were slightly underestimated or overestimated on the day of the Respi-Radar evaluation, with stronger underestimation at epidemiological peaks (Figure 1A, 1B and 1C). The trends were however correctly captured on the day of the evaluation. The incidence of hospital admissions for SARI (from the sentinel network of hospitals) was considerably underestimated between November and December 2023 due to several changes in the surveillance system (expansion of the sentinel network, change in the data collection system). From January 2024 onwards, the situation greatly improved and reflected the trends observed in data from the sentinel network of GPs (Figure 1D). On the other hand, the incidence of SARI hospitalizations with severe complications, collected from the network of hospitals, was never available at the time of the Respi-Radar evaluation. This indicator being collected at patient discharge, typically required four to five weeks to be consolidated (Figure 1E). The number of wastewater treatment plants positive for SARS-CoV-2 was always available at the time of the Respi-Radar evaluation (Figure 1F).

The evolution of each individual indicator was examined in parallel to the overall risk levels attributed over time through the Respi-Radar (Figure 1).

For four indicators of the Respi-Radar (incidence of consultations at GP practices for ILI symptoms; incidence of consultations at GP practices for ARI; incidence of hospital admissions for SARI and incidence of ILI in nursing homes), the evolution taken individually matched the Respi-Radar overall evaluation: increasing trends were observed when the risk level was changed from *green* to *yellow* then from *yellow* to *orange*. Conversely, the trend was decreasing when the risk level of the Respi-Radar changed from *orange* to *yellow* then from *yellow* to *green*. The incidence of complications after hospitalisations for SARI was not available at the time of Respi-Radar analysis, but consolidated data showed a good match with the overall Respi-Radar evaluation.

The indicator on viral concentrations in wastewater did not match the Respi-Radar evaluation, as, during the period considered here (July 2023-March 2024), this indicator remained pathogen-specific, with only SARS-CoV-2 concentrations measured in wastewater while the Respi-Radar evaluated respiratory infections in general.

### b. Feedback from the experts of the RAG

A survey was sent to the RAG experts in April 2024, asking them for feedback on the selection of indicators within the Respi-Radar, the epidemiological thresholds defined for these indicators, the correspondence between the Respi-Radar level and the epidemiological situation, as well as the usefulness of the Respi-Radar as a decision supporting tool.

The participation rate was 69 % (18/26). The indicators and thresholds chosen were generally considered adequate, with the exception of the incidence of severe complications after hospitalization for SARI, for which the delay was considered too long to obtain reliable data. The Respi-Radar was seen by the experts as a useful tool for monitoring trends of respiratory infections in a standardized manner, and for clear communication of the epidemiological situation to the authorities, the healthcare sector and the general public. However, applying a pre-defined set of measures to each determined risk level was challenging, so the decision-supporting aspect of the tool was less recognized.

## Conclusion

The Respi-Radar was implemented at the end of summer 2023, and was used during the autumn-winter 2023-2024 season in Belgium. This season was characterized by an influenza epidemic of moderate intensity and relatively long duration (11 weeks), a COVID-19 peak with limited impact in December 2023, and an RSV peak in October 2023 which intensity was comparable to the previous year. No measures were imposed by the authorities on the healthcare sector or the population, but recommendations were formulated.

The Respi-Radar was built from management tools developed during the COVID-19 crisis to inform and guide political decisions. It aimed at providing an integrated evaluation of the epidemiological situation, taking into account respiratory infections in general, and ensuring a system for early warning.

This unique system in Europe allowed for a standardized evaluation of the epidemiological situation of respiratory infections in Belgium and therefore facilitated communication to authorities, the healthcare sector and even the general population.

The Respi-Radar could be of significant interest to international public health organizations and other countries due to its alignment with the recommendations from the ECDC on respiratory virus surveillance (7). The tool aligns with the ECDC’s call for strengthening surveillance during non-pandemic periods, ensuring responsiveness to emerging viruses and monitoring of multiple respiratory viruses. It also includes both epidemiological and virological data to understand the intensity, spread, and characteristics of viruses. Due to its flexibility, the method could be adopted by countries with differing healthcare infrastructures and surveillance capacities, aiding in building a more resilient international surveillance network.

A limitation, however, is the use of thresholds, which are based on historical data from each surveillance system. In some cases, these data may be incomplete, or system changes may create inconsistencies. Furthermore, thresholds that are effective during non-pandemic periods may not be as useful during pandemics. Even in syndromic surveillance, the introduction of a new pathogen and disruptions to the seasonality of known pathogens can result in unusually high peaks, complicating the detection of changing risks. Therefore, thresholds should be interpreted with caution, requiring expert judgment.

In line with the conclusions drawn during the review of the management tools previously developed for COVID-19 (1), challenges remain to link predefined sets of recommendations to the specific risk levels of the Respi-Radar tool.

The Respi-Radar will be used in the coming respiratory seasons in Belgium. Adaptations are foreseen to improve the detection and monitoring of respiratory infections, but also to establish risk level-based sets of measures. These measures should be implemented in a timely manner to help mitigate and control pressure on the healthcare system.

## Data Availability

All data produced in the present study are available upon request to the authors

## 6- Acknowledgements

The authors sincerely thank the members of the Risk Assessment Group who provided advice and expertise on a regular basis and constitute an essential pillar of this work. The authors also thank all participants to the sentinel surveillances in place in Belgium.

